# Chronomodulated administration of chemotherapy in advanced colorectal cancer. A systematic review and meta-analysis

**DOI:** 10.1101/2022.12.11.22283321

**Authors:** Ahmed Nassar, Amir Abdelhamid, George Ramsay, Mohamed Bekheit

## Abstract

**Aim:** In this systematic review, the efficacy and safety of chronomodulated chemotherapy, defined as delivery of chemotherapy timed according to the human circadian rhythm, were assessed, and compared to continuous infusion chemotherapy for patients with advanced colorectal cancer.

**Methods:** Electronic English-language studies published until October 2020 were searched. Randomised Controlled Trials (RCTs) compared chronomodulated chemotherapy with non-chronomodulated (Conventional) chemotherapy for management of advanced colorectal cancer were included. Main outcomes were the objective response rate (ORR) and system-specific and overall toxicity related to chemotherapy. Electronic database search in Ovid Medline, Ovid Embase, Cochrane CENTRAL and the Cochrane Database of Systematic Review (CDSR).

**Results:** In total, 7 RCTs including 1137 patients were analysed. Males represented 684 (60%) of the study population. The median age was 60.5 (range: 47.2 - 64) years.

There is no significant difference between chronomodulated and conventional chemotherapy in ORR ((risk ratio (RR) 1.15; 95%CI [0.87-1.53]). There was no significant difference in gastrointestinal toxicity under the random effect model RR 1.02; CI 95% [0.68 – 1.51]. No significant difference was found regarding neurological and skin toxicities, (RR 0.64,95%CI [0.32 – 1.27]) and (RR 2.11, 95%CI [0.33-13.32]), respectively. However, patients who received chronomodulated chemotherapy had fewer haematological toxicity, (RR 0.36, 95%CI [0.27-0.48]).

**Conclusion:** There was no overall difference in ORR or toxicity but haematologic, between chronomodulated and non-chronomodulated chemotherapy used for patients with advanced colorectal cancer. Chronomodulated chemotherapy could be considered in patients at high risk of haematological toxicities.

## Background

Circadian rhythm is based on two main mechanisms. The first mechanism is a central system which acts coordinator and includes suprachiasmatic nuclei located in the hypothalamus and considered as the main circadian pacemaker.^1^ The second mechanism is a molecular “clock” which is present in most cells in brain and peripheral tissue and consists of multiple feedback loops produced by transcriptional and post-transcriptional process triggered by genes responsible of specific proteins expression in rhythmic manner.^1,2^ Effects on cell division cycle related changes such as apoptosis and cell repair paves the way to investigate developing cancer chemotherapeutic regimens.^1,3^

The circadian rhythm plays a role in several biological processes. At least 15 specific genes are believed to be related to circadian rhythm taking part in controlling cell proliferation, DNA replication, apoptosis, angiogenesis, metabolism, and drug detoxification.^4,5^ There is an observed 24 hours change in the activities of several enzymes involved in catabolism of different chemotherapeutic factors or anabolism of its cytotoxic forms.^6–8^ Hence, circadian rhythm could be related to the efficacy of cancer treatment under what is known as chronotherapy, which is an approach that could potentially improve the tolerability and efficacy of chemotherapy. ^5^

Chronomodulation of chemotherapy is based on utilising the circadian rhythm to increase the efficacy of anti-neoplastic agents.^9^ It was found that some isoforms of heat shock protein HSP90, which mediate cell-cycle progression, shows circadian pattern of expression and this may explain circadian rhythm dependent efficacy of some anti-cancer agents.^10,11^ Moreover, toxic effects of endotoxins^12^ and anti-cancer agent, Cyclophosphamide, show dependency on time of the day.^13,14^

Over the last two decades, several experimental and clinical studies have shown a favourable association between adjusting timing and dosing of chemotherapeutic agents according to circadian rhythm and response in cancer patients.^3^ L-Alanosine is an amino acid analogue derived from bacterium Streptomyces alanosinicus and shows antimetabolic and potential anti-neoplastic activity.^15^ It has shown a selective in vitro anti-neoplastic activity against Methylthioadenosine phosphorylase (MTAP) deficient tumours ^1,16^ such as, leukemias, brain tumours, non–small-cell lung cancers, breast cancers, melanomas, pancreatic cancers, and sarcomas.^17–21^ However, bone marrow suppression and mucositis are common causes of dose limitation and discontinuation.^1,22^ One study in mice has proven three-folds decrease in mortality with circadian rhythm adjusted doses of L-Alanosine, confirming a potential strong role of circadian rhythm. ^1^ Chronomodulation of chemotherapy has yet to become standard practice in other disease settings and it remains unclear if such an approach may influence outcomes.

Colorectal cancer is the 4^th^ most common cancer in the United Kingdom (UK), accounting for 11% of all new cancer cases.^23^ It is the second most common fatal cancer after lung cancer in western countries.^6,24^ Metastases are detected in 25% to 30% patients at the time of diagnosis and will develop during the disease course in further 25% of patients^6^ Metastases are responsible for 90% of deaths from colorectal cancer.^25^ The liver is the most common organ site for metastatic disease.^26^ In these advanced cases, chemotherapy is indicated for control of the systemic disease, which may not be controlled only by surgery.^4,6,27^

As a result of the rate of advanced presentation, and rates of metastatic disease, chemotherapy regimens are commonplace in colorectal cancer practice. However, the optimal mechanism by which such therapy is delivered remains unclear. Fluorouracil (5 FU) and leucovorin (folinic acid) (LV) are included in most chemotherapy regimens for colorectal cancer and result in an objective response (i.e. decrease in tumour size by 50% or more) in 20-25% of patients and up to 50% if combined with other agents like Oxaliplatin with a dose-related response. ^5,28,29^ This indicates that if the chemotherapy is tolerated well, the patient may benefit from the full therapeutic dose^29^. However, between 31% to 34% of patients, experience severe toxicity from 5 FU.^30^ Myelosuppression resulting in severe neutropenia and anaemia is the main toxicity of 5 FU.^31^ Gastrointestinal toxicity resulting in diarrhoea and mucositis can happen but is less frequent.^31^ Likewise, Oxaliplatin is associated with anaemia which can be severe if combined with 5 FU.^32^ Pre-operative anaemia in colorectal cancer is associated with poor disease progression and post-operative recovery.^33^ Additionally, Oxaliplatin is associated with neurotoxicity and fatigue.^34^

Chronomodulation of chemotherapy is defined as delivery of chemotherapy in respect to the circadian rhythm in which different doses of chemotherapy will be delivered according to the time of the day.^24,29,35^ In this approach, circadian rhythm related changes can be utilised to improve tolerance and efficacy of chemotherapy.^25,36^ There are, however, controversies regarding the tolerability and efficacy of chronomodulated chemotherapy regimen compared to the conventional (non-chronomodulated) regimen despite the presence of randomised controlled trials comparing these two approaches of chemotherapy for advanced colorectal cancer .^25^

Despite studies have shown a potential beneficial role of chronomodulation as previously mentioned, its effect on objective response rate (ORR) and different body systems toxicities are yet to be proven. Other factors that may affect results of chronomodulation still need to be explored, too. Hence, recent synthesis of the currently published literature in this field has yet to be undertaken. In this review, the efficacy and safety of chronomodulated chemotherapy were assessed and compared to continuous infusion chemotherapy for patients with advanced colorectal cancer.

## Methods

### Study design

This review was prepared in line with the Preferred Reporting Items for Systematic reviews and Meta-analyses (PRISMA) statement. This systematic review was registered to PROSPERO (University of York) before study selection process (registration number: CRD42020227313)

### Inclusion criteria

Electronic English-language studies published until October 2020 were searched. Randomised Controlled Trials (RCTs) comparing chronomodulated chemotherapy with conventional chemotherapy for the management of advanced colorectal cancer were included. The main outcomes were the objective response rate (ORR) and system-specific and overall toxicity related to chemotherapy.

### Exclusion criteria

Observational studies, reviews and non-randomised controlled studies were excluded. Study was to be excluded if included patients with non-colorectal cancer.

### Search Strategy

Electronic database search in Ovid Medline, Ovid Embase, Cochrane CENTRAL and the Cochrane Database of Systematic Review (CDSR). The search was conducted by a senior information specialist from the library department of the Royal College of Surgeons of England and was executed on the 27^th^ of October 2020.

### The study question

In patients with advanced colorectal cancer, what is the effect of chronomodulated chemotherapy compared to conventional chemotherapy on ORR and chemotherapy related toxicities? Patient Intervention Control Outcome (PICO) framework was used to guide the search (supplementary table 1). Full electronic search strategy is shown in supplementary table 2.

Two independent blinded reviewers performed the abstract screening. Any conflicts were resolved by a third reviewer to produce the final list of studies eligible for full-text review. Full text review and data extraction from individual studies were carried out by two researchers with another researcher to confirm adequacy and accuracy of data extracted. Data included each study details, demographic data, details of chemotherapy regimen, disease characteristics, previous treatment (chemotherapy, radiotherapy, and surgery), objective response rate and specific toxicities in both treatment arms. Follow up period and withdrawals from each study were also noted. The revised Cochrane risk-of-bias tool for RCTs (RoB2) Tool was used to assess risk of bias from all included RCTs.^37^

### Definitions

The Control group had conventional (non-chronomodulated) chemotherapy and was referred to as control group group A) while treatment group had chronomodulated chemotherapy and was referred to as treatment group (group B).

The Objective response rate (ORR) is the assessment of the tumour burden after a given treatment and it was measured according to WHO criteria for disease response.^38^

The term toxicity refers to toxicity secondary to chemotherapy involving one or more of the gastrointestinal, haematological, neurological, and dermatological systems.

Toxicity was graded according to National Cancer Institute-Common Toxicity Criteria.^39^ (supplementary table 3)

‐ The chemotherapeutic agent utilized in the included studies were: 5-FU: Fluorouracil, LV: Leucovorin (folinic acid), CPT-11: Irinotecan, FUDR: Floxuridine and I-OHP: Oxalatoplatinum
‐ As different regimens were included in the studies this review relied on, these regimens of chemotherapy are defined here as follows:

Regimen 1: 5-FU, LV plus oxaliplatin

Regimen 2: intra-arterial 5 FU, Oxaliplatin

Regimen 3: irinotecan CPT-11, 5-FU, LV

Regimen 4: venous 5-FU + arterial FUDR

Regimen 5: 5-FU, I-OHP and LV

### Data handling

Only grade 3 and 4 toxicities were analysed as they can be a cause of interrupting chemotherapy course or reducing dose due non-tolerability.

### Statistical analysis

Count, percentages, and ratios were used to represent categorical variables and median (range) was used to represent continuous data as stated in each individual study. Outcomes, such as ORR and toxicity, were represented by risk ratios (RR) and 95% confidence interval (CI).

Meta-analysis was conducted using RevMan (Review Manager) software version 5.4 (Cochrane collaboration, United Kingdom). Analysis was done for different types of toxicities. I^2^ and Tau^2^ values were used to assess heterogeneity. If the I^2^ value was > 50%, significant heterogeneity was considered and Mantel-Haenszel (M-H) random effect model to be employed.^40^ Meta-regression was performed using Comprehensive Meta-analysis software version 3. Significant difference was considered if P-value was of less than Funnel plots were produced to visualize risk of publication bias across studies and significant asymmetry was an indication of publication bias.

### Ethical review

This was a meta-analysis of data already published in RCTs and thus ethical review was not required.

## Results

### Studies Characteristics

Out of 260 studies identified in the searches, 70 were duplicates. After screening and eligibility check, 7 RCTs were included in quantitative analysis. Figure 1 displays the selection stratification (in PRISMA format). All included RCTs were of parallel randomised design except Levi et al’s 1997 was designed as a cluster-randomised trial.^29^ All trials included patients with advanced colorectal cancer with or without metastasis and needed chemotherapy. Five trials were multicentre studies. ^29,35,41–43^ The main study characteristics, including inclusion and exclusion criteria, are summarised in table 1. There were differences in the inclusion and exclusion criteria of eligible patients across studies. In one study, Ramanathan et al, 4 treatment arms were compared.^41^ All of them received the same regimen (regimen 1) but different timings except in arm 1 where leucovorin (LV) was not included. Hence, arm 1 (n=23) was excluded from the meta-analysis. Arms 2 and 3 were considered as the group A (conventional chemotherapy) (n=81) and arm 4 was the group B (chronomodulated chemotherapy) (n=25).

**Table 1:**
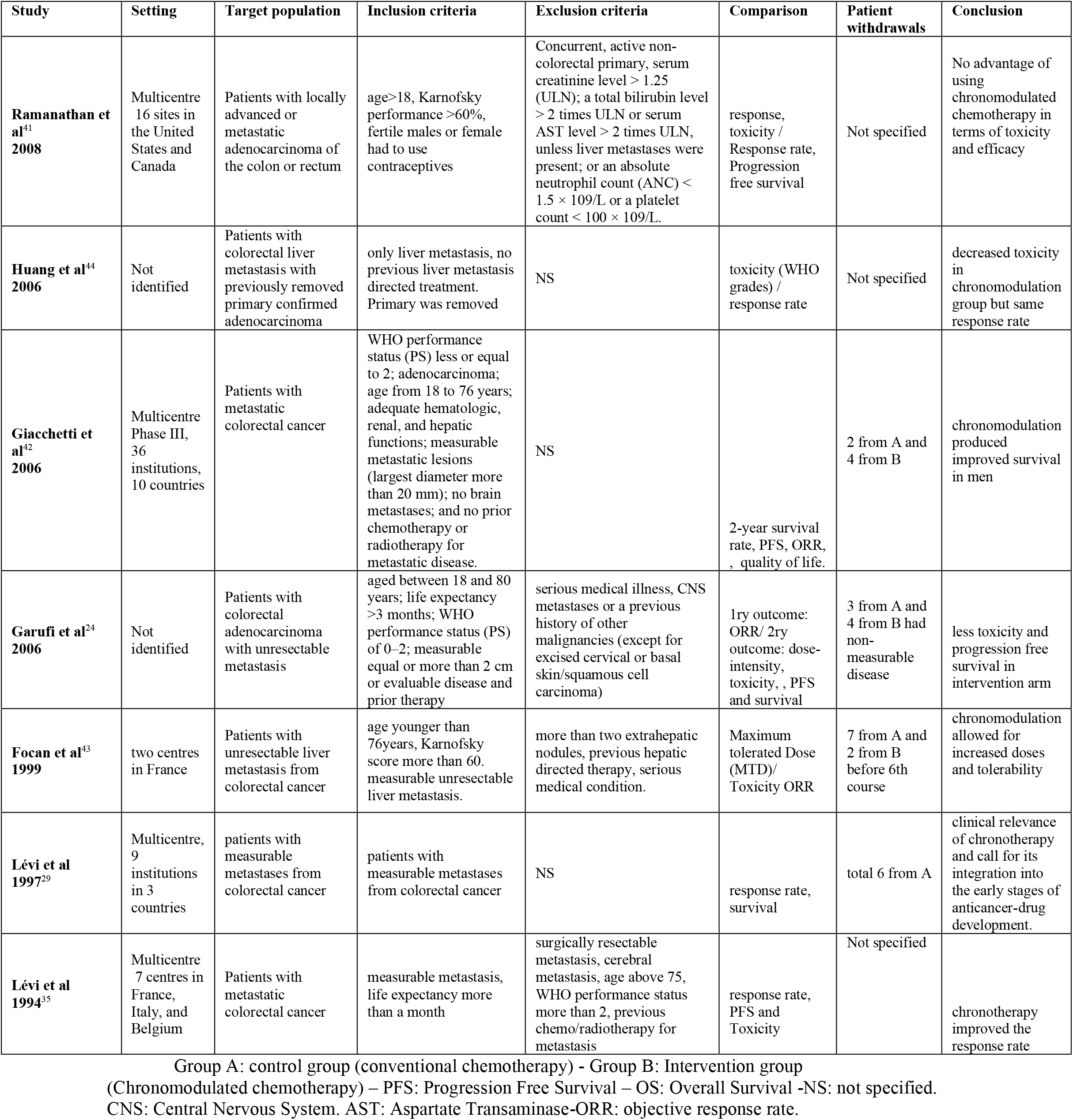
Studies characteristics

| Study | Setting | Target population | Inclusion criteria | Exclusion criteria | Comparison | Patient withdrawals | Conclusion |
| --- | --- | --- | --- | --- | --- | --- | --- |
| **Ramanathan et al<sup>41</sup> 2008** | Multicentre<br>16 sites in the United States and Canada | Patients with locally advanced or metastatic adenocarcinoma of the colon or rectum | age>18, Karnofsky performance >60%, fertile males or female had to use contraceptives | Concurrent, active non-colorectal primary, serum creatinine level > 1.25 (ULN); a total bilirubin level > 2 times ULN or serum AST level > 2 times ULN, unless liver metastases were present; or an absolute neutrophil count (ANC) < 1.5 × 10<sup>9</sup>/L or a platelet count < 100 × 10<sup>9</sup>/L. | response, toxicity / Response rate, Progression free survival | Not specified | No advantage of using chronomodulated chemotherapy in terms of toxicity and efficacy |
| **Huang et al<sup>44</sup> 2006** | Not identified | Patients with colorectal liver metastasis with previously removed primary confirmed adenocarcinoma | only liver metastasis, no previous liver metastasis directed treatment. Primary was removed | NS | toxicity (WHO grades) / response rate | Not specified | decreased toxicity in chronomodulation group but same response rate |
| **Giacchetti et al<sup>42</sup> 2006** | Multicentre Phase III, 36 institutions, 10 countries | Patients with metastatic colorectal cancer | WHO performance status (PS) less or equal to 2; adenocarcinoma; age from 18 to 76 years; adequate hematologic, renal, and hepatic functions; measurable metastatic lesions (largest diameter more than 20 mm); no brain metastases; and no prior chemotherapy or radiotherapy for metastatic disease. | NS | 2-year survival rate, PFS, ORR, , quality of life. | 2 from A and 4 from B | chronomodulation produced improved survival in men |
| **Garufi et al<sup>24</sup> 2006** | Not identified | Patients with colorectal adenocarcinoma with unresectable metastasis | aged between 18 and 80 years; life expectancy >3 months; WHO performance status (PS) of 0–2; measurable equal or more than 2 cm or evaluable disease and prior therapy | serious medical illness, CNS metastases or a previous history of other malignancies (except for excised cervical or basal skin/squamous cell carcinoma) | 1ry outcome: ORR/ 2ry outcome: dose-intensity, toxicity, , PFS and survival | 3 from A and 4 from B had non-measurable disease | less toxicity and progression free survival in intervention arm |
| **Focan et al<sup>43</sup> 1999** | two centres in France | Patients with unresectable liver metastasis from colorectal cancer | age younger than 76years, Karnofsky score more than 60. measurable unresectable liver metastasis. | more than two extrahepatic nodules, previous hepatic directed therapy, serious medical condition. | Maximum tolerated Dose (MTD)/ Toxicity ORR | 7 from A and 2 from B before 6th course | chronomodulation allowed for increased doses and tolerability |
| **Lévi et al 1997<sup>29</sup>** | Multicentre, 9 institutions in 3 countries | patients with measurable metastases from colorectal cancer | patients with measurable metastases from colorectal cancer | NS | response rate, survival | total 6 from A | clinical relevance of chronotherapy and call for its integration into the early stages of anticancer-drug development. |
| **Lévi et al 1994<sup>35</sup>** | Multicentre 7 centres in France, Italy, and Belgium | Patients with metastatic colorectal cancer | measurable metastasis, life expectancy more than a month | surgically resectable metastasis, cerebral metastasis, age above 75, WHO performance status more than 2, previous chemo/radiotherapy for metastasis | response rate, PFS and Toxicity | Not specified | chronotherapy improved the response rate |

Group A: control group (conventional chemotherapy) - Group B: Intervention group (Chronomodulated chemotherapy) – PFS: Progression Free Survival – OS: Overall Survival -NS: not specified. CNS: Central Nervous System. AST: Aspartate Transaminase-ORR: objective response rate.

**Figure 1:**
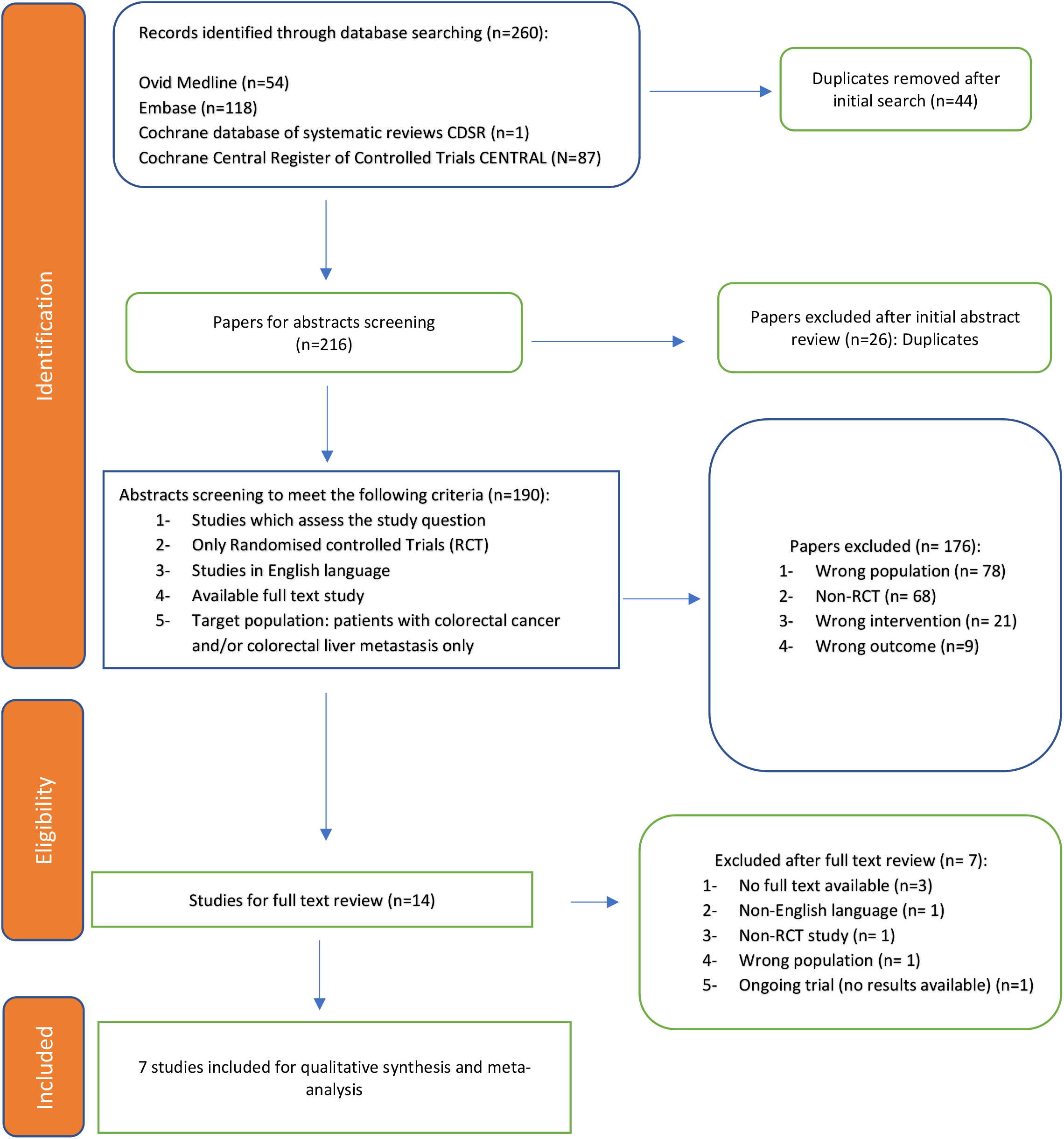
PRISMA diagram

**Figure 1:**
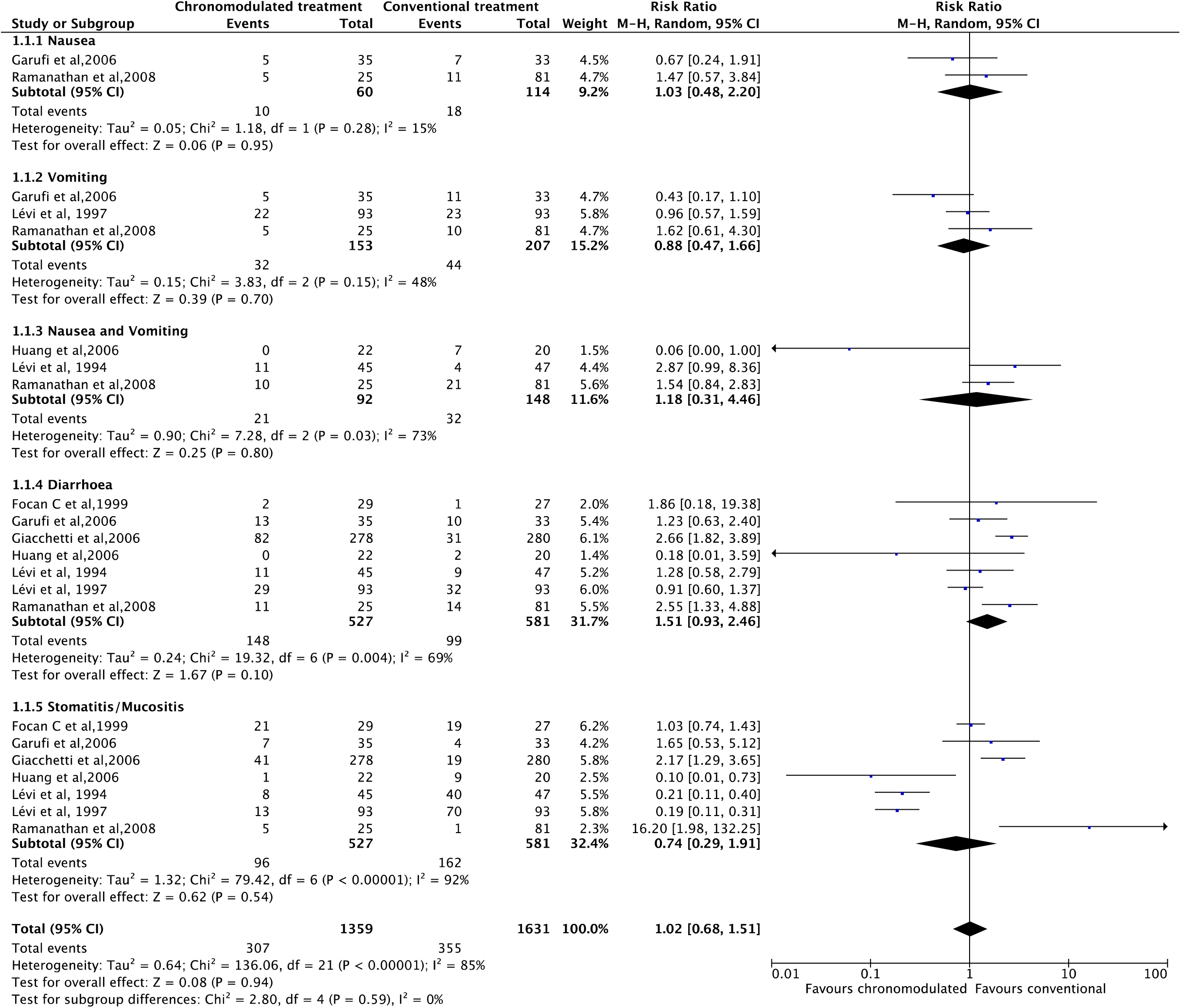
Grade 3 and 4 gastrointestinal toxicities; Chronomodulated vs conventional chemotherapy

### Risk of bias and quality assessment

One trial was of high risk of bias^43^, four trials were with some concerns^35,41,42,44^, and two were of low risk of bias according to ROB2 assessment tool^24,29^. Further details are available on supplementary table 4.

The number of patient withdrawals from each trial was noted as 18 patients from conventional treatment group and 10 patients from the chronomodulated group from 4 studies^24,29,42,43^ Number of withdrawals was not specified in the other three studies.^35,41,44^

There was no significant risk of bias regarding ORR and haematological toxicity (supplementary figures: 1 and 2). However, there was a significant risk of bias in gastrointestinal toxicity (supplementary figure:3). There was not enough number of studies to assess risk of publication bias in other types of toxicities.

### Patient characteristics

In total, out from 7 RCTs, 1137 patients were included. Males represented 684 (60%) of the study population. The median age was 60.5 (range: 47.2 – 64) years. The majority of patients (90%) were of 0 and 1 WHO performance status in two studies.^24,42^ Karnofsky performance score was more than 60% in other two studies.^41,43^ There was no reported significant difference in patients’ characteristics between the two treatment groups within each individual study. A detailed description of the patient characteristics are listed in supplementary table 5.

### Disease characteristics

The colon was the primary site in 842 (74%) patients compared to rectum which was the primary site in 295 (26%) patients. In 4 studies, 418 (46%) of patients had metastasis in 2 or more sites.^24,29,35,42^ In same studies, the liver was involved in 757(83%) patients and lung was involved in 322 (35%) patients.^24,29,35,42^ In Focan et al 45 (80%) patients had isolated liver involvement.^43^ Haung et al has included patients with only liver metastasis and 28 (66.7%) patients had both lobes involved.^44^ 455 (73%) patients were staged initially as Duke’s stage D (synchronous metastasis) in two studies.^42,43^ 162 (16.7%) had previous chemotherapy and 74 (7.6%) had previous radiotherapy in 5 studies.^24,29,35,42,43^ Focan et al reported significant discrepancy between both treatment groups regarding patients who had previous therapy (chemotherapy, radiotherapy or combined) where 6 (22%) patients had prior therapy in group A compared with 15 (52%) patients in group B. ^43^ (supplementary table 6)

### Chemotherapy Regimen

All chemotherapeutic agents were delivered through a programmable pump through intravenous (IV) access except two trials (Huang et al and Focan et al) where an access to the hepatic artery was established prior commencing the first course. Chemotherapeutic agents used were similar in Ramanathan et al and Giacchetti et al. 5-Fluorouracil (5-FU), Leucovorin (LV) and Oxaliplatin were used in both studies (regimen 1). In Garufi et al, chronomodulated 5-FU and leucovorin (folinic acid) (LV) were administered in both treatment groups but chronomodulated irinotecan was given in the intervention group only and this compared (regimen 3). Chronomodulation of specific agent was arranged to ensure peak flow at either 4:00h or 16:00h if a second chronomodulated agent administered. (Supplementary table 7)

### Objective response rate (ORR)

In Ramanathan et al, ORR was measured at weeks 6,12,18 after start of treatment and at day 28 post treatment.^41^ Two trials (Huang et al and Giacchetti et al) assessed ORR after two and 4 courses respectively.^42,44^ Three trials measured ORR every 3^rd^ course.^29,35,43^

Meta-analysis was conducted using data from all studies. Under the random effect model, there is no significant difference between chronomodulated and conventional chemotherapy for ORR (RR 1.15; 95% CI [0.87-1.53]). (Figure 2)

**Figure 2:**
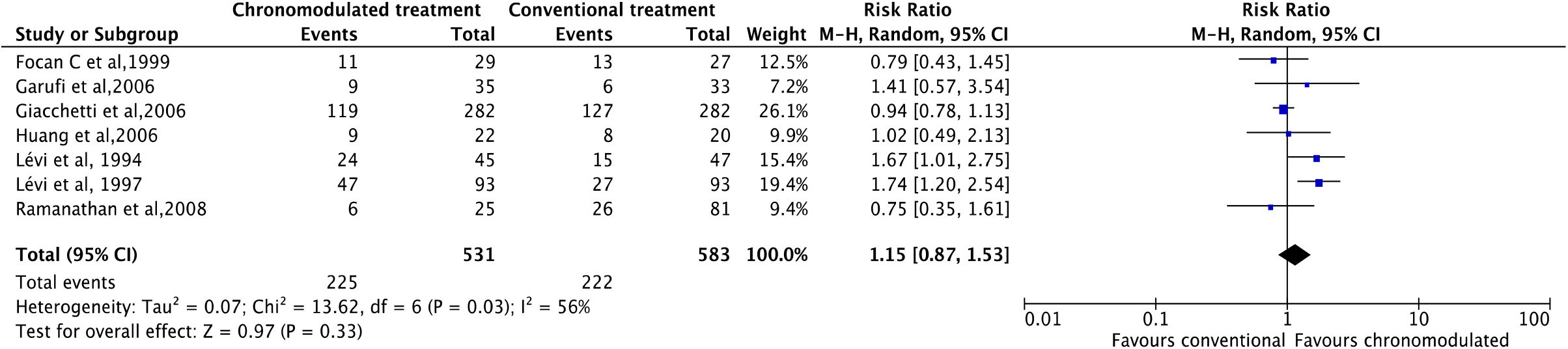
Objective response rate (ORR); chronomodulated vs conventional chemotherapy

### Toxicity

Grades 3 and 4 of toxicity were assessed. Toxicity was measured for 4 main systems: (Gastrointestinal, Haematological, Neurological, and Skin). There was no significant difference in gastrointestinal toxicity under the random effect model (RR=1.02, CI 95% [0.68 – 1.51]) (figure:3).

However, the chronomodulated arm had a 63% less chance of developing haematological toxicity, (RR 0.36, 95%CI [0.27-0.48]). (Figure 4) Patients received chronomodulated chemotherapy had similar neurological toxicities compared to conventional treatment (figure: 5), (RR 0.64, 95%CI [0.32 – 1.27]). Similarly, there was no significant difference between both group regarding skin toxicities, (RR 2.11,95%CI [0.33-13.32]), (figure: 6).

**Figure 4:**
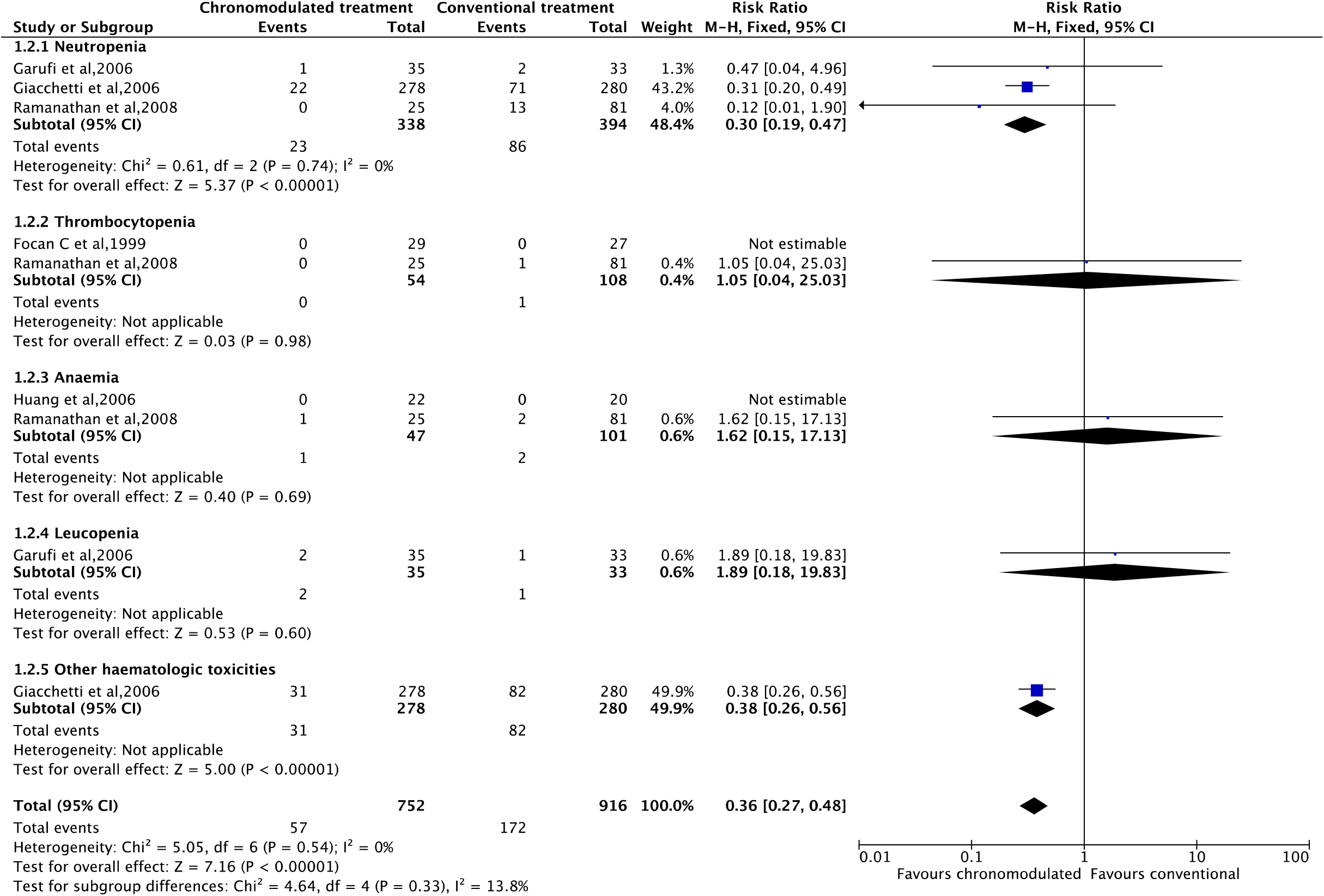
Grade 3 and 4 haematological Toxicities; Chronomodulated vs conventional chemotherapy

**Figure 5:**
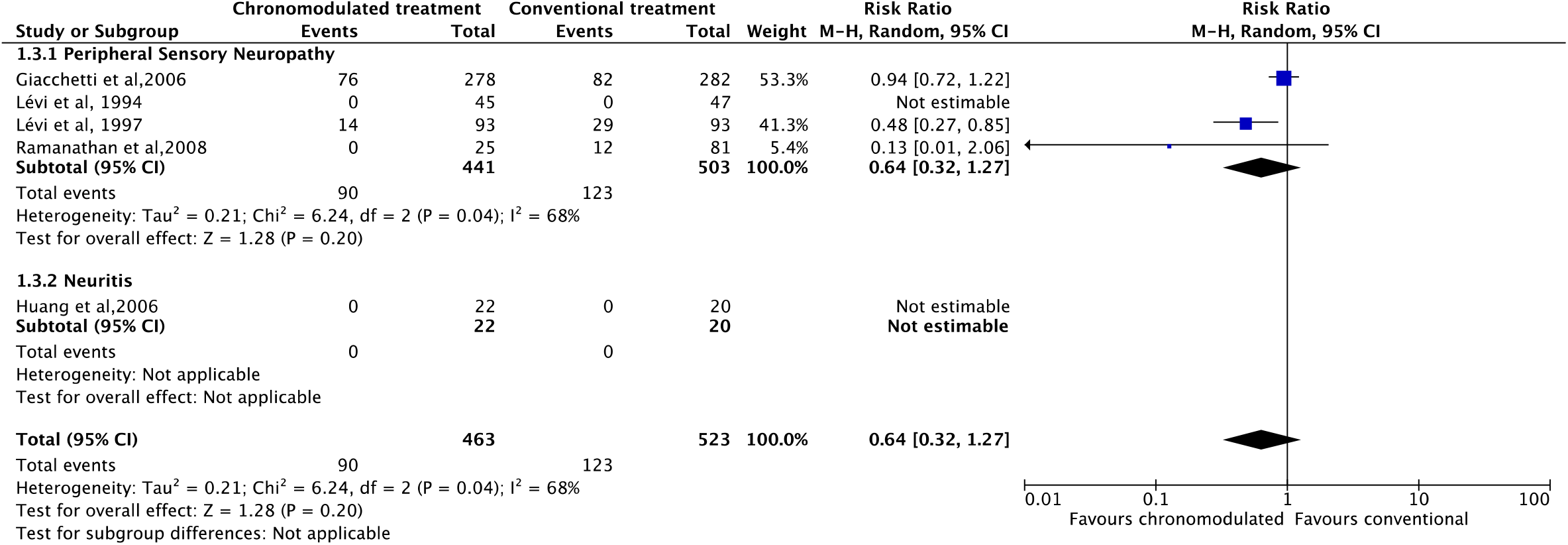
Grade 3 and 4 neurological Toxicities; Chronomodulated vs conventional chemotherapy

**Figure 6:**
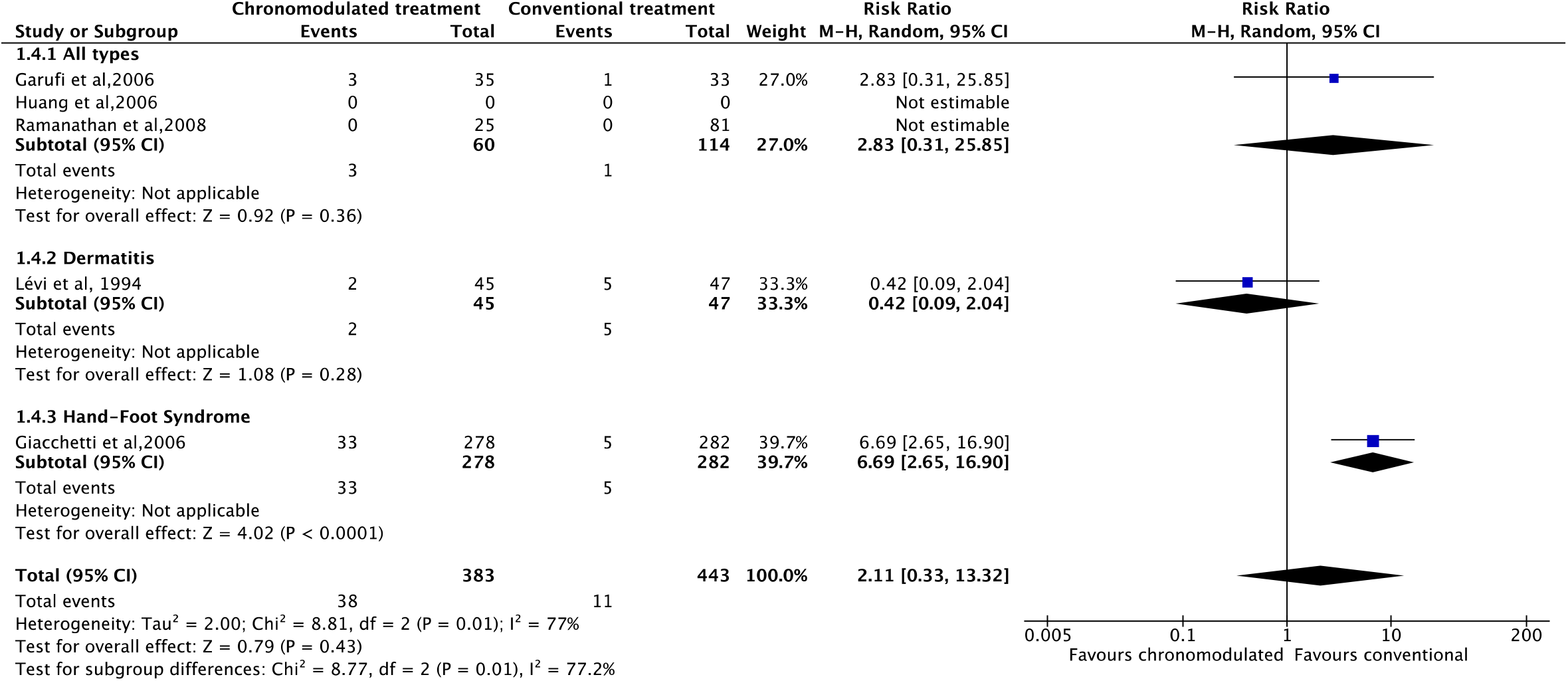
Grade 3 and 4 skin Toxicities; Chronomodulated vs conventional chemotherapy

The overall risk of grade 3 and 4 toxicities was not different between both groups (RR 1.00, CI [0.57-1.75]). Meta-Regression (Supplementary Table 8 and supplementary figure 4) showed that chemotherapy regimen 2 (i.e., intra-arterial 5-FU and oxaliplatin) had a significantly less risk of toxicity (p = 0.0048). Of note, one study reported the incidence of grade 3 or 4 main toxicities was greater by 15.3%, (95% CI [7.5-23.2]) in women compared to men from the chronomodulated treatment group. ^42^

## Discussion

In this meta-analysis of 7 RCTs, there was not a significant advantage of chronomodulated chemotherapy in improving the response rate and gastrointestinal, neurological, and skin toxicities. However, haematological toxicity was significantly lower compared to conventional regimen.

Liao et al, assessed the overall survival and safety were patients on chronomodulated chemotherapy were more prone to diarrhoea but at less risk of neutropenia. There was no difference in overall survival and response rate.^6^ In contrast with Liao et al study in which intra-venous administration was the only method of delivery, this review included two extra RCTs compared chronomodulated and conventional intra-arterial administration of chemotherapeutic agents. Moreover, effect of different chemotherapeutic regimens on toxicity were studied in meta-regression which suggested better tolerance of intrahepatic artery versus systemic delivery of 5 FU.

There were several advantages for patients on chronomodulated regimen. They were 3 times less likely to develop neutropenia and developed fewer stomatitis compared to conventional regimen. Furthermore, the number of withdrawals was significantly higher in the non-chronomodulated group due to either severe toxicity or disease progression ^29,42^.

Giacchetti et al, included the largest number of patients in their trial (564 patients) and reported a significantly (15.3%, 95%CI =7.5-23.2) greater incidence of grade 3 and 4 toxicity incidence in women ^42^. However, the overall mortality was higher in women than in men (38% vs 25%, P<0.01) and the same effect was noticed on progression free survival ^42^. This may suggest a strong role of gender on the efficacy of chronomodulation and paves the way for future studies.

Intra hepatic-arterial administration of chemotherapy for colorectal liver metastasis was reported to be more effective than systematic administration.^45^ Meta-regression found significantly lower risk of toxicity in patients who had intra-arterial 5-FU and oxaliplatin. However, this was not the case for patients who had intra-arterial FUDR and venous 5-FU, suggesting more tolerability for 5-FU if given intra-arterial rather than systemic. The influence of previous chemotherapy on the effect of chrnomodulation is not clear. Focan et al, reported a significantly larger number of patients with previous chemotherapy in the chronomodulation group. This may have masked the difference in ORR and toxicity reported in other trials in which they had matched distribution of number of patients who had previous chemotherapy among both groups.^43^

The activity of many enzymes regulating the anabolism and catabolism of agents like 5-FU and oxaliplatin has shown a circadian variation.^11,32^ At least 50% of the proportion of cells in the S-phase were shown to change during the day.^47^ Therefore, circadian rhythms can alter the tolerability of patients to chemotherapy and improve its antitumor efficacy when administrated near their respective times of best tolerability.^25^ Time of best tolerability and efficacy depends on circadian changes of enzymes involved in metabolism of each agent.^7,46^ This was the predominant reason behind the scheduled chronomodulated chemotherapy at different peak doses considering the chemotherapeutic agent (e.g., peak dose at 4:00 a.m for 5-FU and at 4:00 p.m for oxaliplatin).^5,29^

This review has some limitations. According to the risk of bias assessment (supplementary table 4), there were some concerns in the majority of studies and high concern of bias in one study. There was also, a considerable heterogeneity in the meta-analysis which is not explained by the meta-regression for different chemotherapeutic agents as R^2^ value was 0 (supplementary table 8), therefore, random effects model was used. Potential confounding factors such as previous chemotherapy and the volume and location of distant metastasis were not reported by all studies. All included RCTs calculated their sample size based on ORR as a primary outcome and it may not be sufficient to study toxicity. Further studies needed to address the effect of gender and other disease specific factors on chronomodulation. Age and disease-stage-specific characteristics should be taken into consideration to explore its impact on usefulness of this approach to improve the outcome for patients who will benefit from chronomodulated chemotherapy with advanced colorectal cancer. In addition, some of secondary outcome’s comparisons such as neutropenia, were based only on limited number of studies. Despite meta-regression was conducted to examine the effect of different chemotherapeutic regimens, this may be influenced by the underlying heterogeneity of disease stage. Hence, future studies need to consider disease stage among other disease and patients characteristics. This consideration may allow better assessment of influence of the different chemotherapeutic regimens and their delivery approach.

## Conclusion

There was no difference in ORR and overall toxicity between chronomodulated and non-chronomodulated chemotherapy used in patients with advanced colorectal cancer. Chronomodulated chemotherapy could be considered in patients at high risk of developing haematological toxicities. chronomodulation may be more tolerable in men. Further high-quality studies are recommended to ascertain the current findings.

## Data Availability

All data produced are available online at: https://1drv.ms/u/s!AlzdopwEri123kc7F0pt4YnN8drW?e=lylmBq

## List of abbreviations

5 FU: Fluorouracil
AST: Aspartate Transaminase
CDSR: Cochrane Database of Systematic Review
CI: Confidence Interval
CNS: Central Nervous System
CPT-11: Irinotecan
DNA: Deoxyribonucleic acid
FUDR: Floxuridine
I-OHP: Oxalatoplatinum
LV: leucovorin
M-H: Mantel-Haenszel
MTAP: Methylthioadenosine phosphorylase
NS: not specified
ORR: objective response rate
OS: Overall Survival
PFS: Progression Free Survival
PICO: Patient Intervention Control Outcome
PRISMA: Preferred Reporting Items for Systematic reviews and Meta-analyses
RCTs: Randomised Controlled Trials
RevMan: Review Manager
RoB2: Risk of Bias 2 tool
RR: Risk ratio
WHO: World Health Organisation

## Declarations

### Competing interests

The authors declare that they have no competing interests

### Ethics approval and consent to participate

Not applicable

### Consent for publication

Not applicable

### Availability of data and materials

The datasets generated during and/or analysed during the current study are available in the [SoF copy2.xlsx] and [chronomodulation.rm5] at : https://1drv.ms/u/s!AlzdopwEri123kc7F0pt4YnN8drW?e=lylmBq

### Funding

No funding

### Authors’ contributions

**AN:** contributed to abstract screening, full text review, data extraction, analysis, designing and writing up manuscript. **AA:** contributed to abstract screening, full text review and data extraction. **GR:** contributed to substantive and major revision of the draft. **MB:** contributed to conceptualisation, resolution of conflicts during abstract screening and full text review, major revision of the draft and is the corresponding author. All authors approved the final version prior to submission.

## Acknowledgements

With thanks to Kirsty Morrison, Senior Information Specialist, Royal College of Surgeons of England Library and Archives Team, for conducting the literature searches

## Supplementary Material

**supplementary table 1:** PICO Framework 1

| PICO | Description | Search Terms |
| --- | --- | --- |
| **Population** | Patients diagnosed with colorectal cancer and/or colorectal liver metastasis cancer undergoing chemotherapy. | Gastrointestinal Neoplasms/ or Liver Neoplasms/ or Carcinoma, Hepatocellular/ or Biliary Tract Neoplasms/ or Bile Duct Neoplasms/ or Cholangiocarcinoma/ digestive system cancer/ or biliary tract tumor/ or bile duct carcinoma/ or liver tumor/ or hepatobiliary system cancer/ or liver cell carcinoma/ or gallbladder cancer/colon cancer/ rectal cancer/ |
| **Intervention** | Chrono-modulated chemotherapy | Chrono-modulated chemotherapy, chronotherapy, chronomodulated chemotherapy, chronomodulation |
| **Control** | Non-chronomodulated chemotherapy /standard chemotherapy/ Conventional chemotherapy | Drug Therapy/ or Antineoplastic Agents/Chemotherapy/ Chronomodulated chemotherapy/Chronotherapy |
| **Outcome** | Objective response rate (ORR), toxicity , associated symptoms during chemotherapy (e.g., vomiting, nausea, headache, etc.) | No search terms used to find more results |

**Supplementary table 2:**
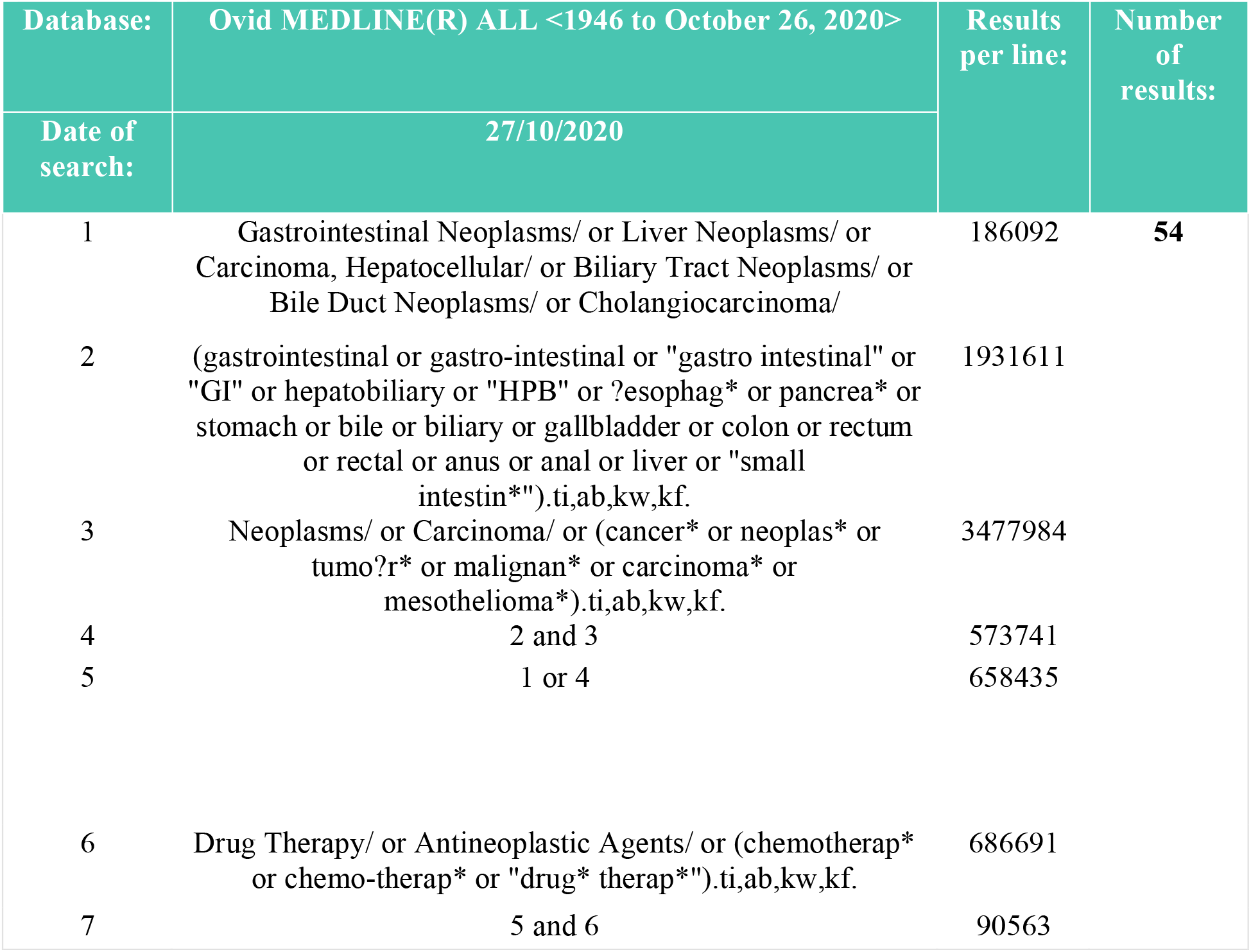
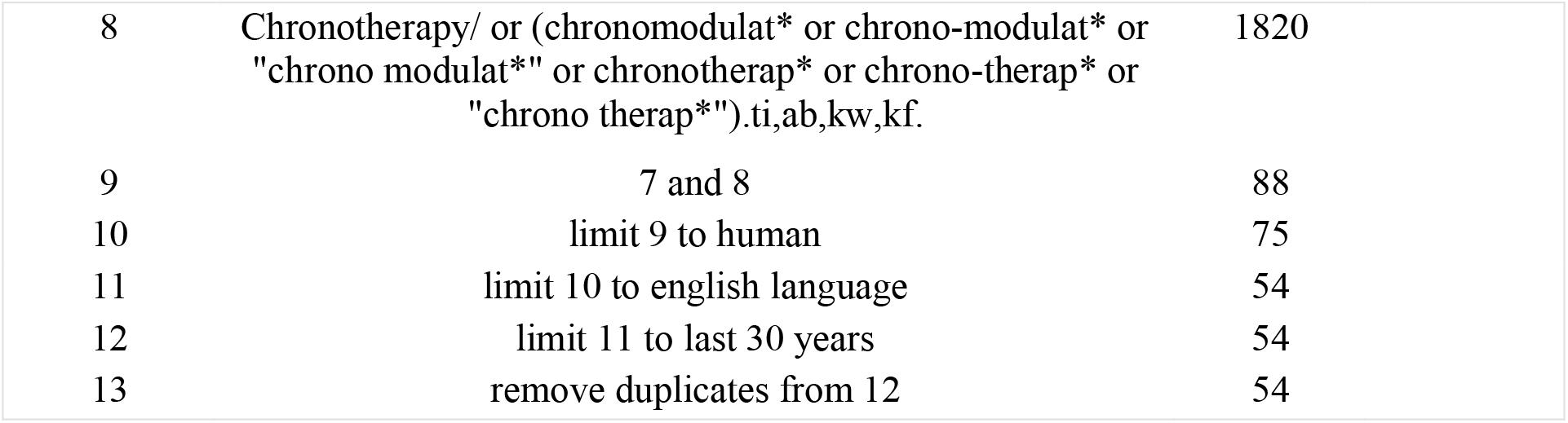
Search in Ovid MEDLINE database

**Supplementary table 3:** Grading of toxicity

| Grade of toxicity | Explanation |
| --- | --- |
| Grade 1 | Mild; asymptomatic or mild symptoms; clinical or diagnostic observations only; intervention not indicated. |
| Grade 2 | Moderate; minimal, local, or non-invasive intervention indicated |
| Grade 3 | Severe or medically significant but not immediately life-threatening; hospitalization or prolongation of hospitalization indicated |
| Grade 4 | Life-threatening consequences: urgent intervention indicated. |

**Supplementary table 4:**
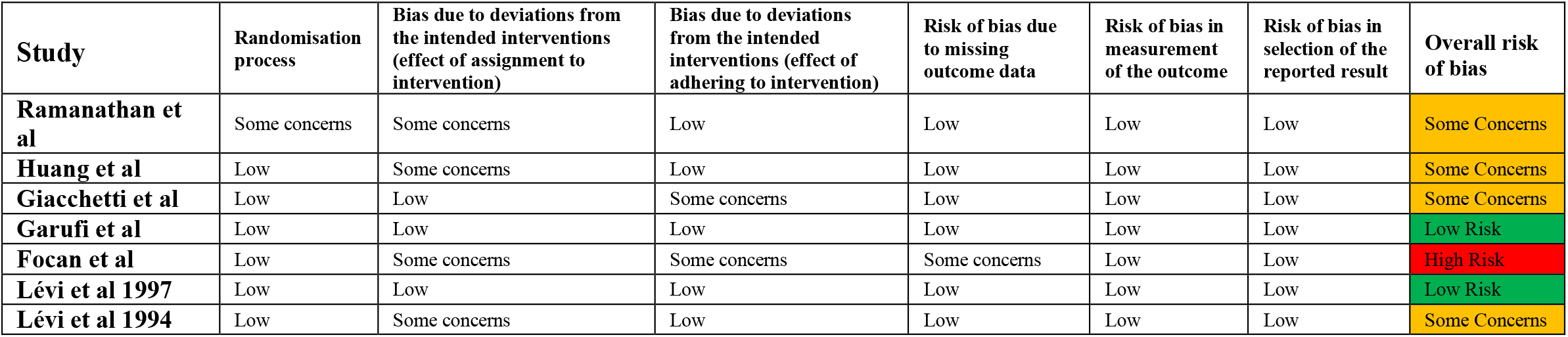
Assessment of risk of bias (RoB2 tool)

**Supplementary table 5:** Patients Characteristics:

| Study | Sample size | Group A | Group B | Age (Median-Range) |  | Gender (M: F) |  | Performance status (WHO) |  |
| --- | --- | --- | --- | --- | --- | --- | --- | --- | --- |
|  |  |  |  | Group A | Group B | Group A | Group B | Group A | Group B |
| **Ramanathan et al 2008** | 129 | 104 | 25 | **2 arms:** 60 (27-78) and 55(25-75) | 61(43:81) | 59:22<br>72.8% males | 18:7<br>72% males | 0: 55 (67.9%)<br>1: 25 (30.8%)<br>2: 1 (1%) | 0: 15 (60%)<br>1: 10 (40%)<br>2: 0 |
| **Huang et al 2006** | 42 | 20 | 22 | 47.2 | 53.4 | 8:12<br>40% males | 11:11<br>50% males | Not specified | Not specified |
| **Giacchetti et al 2006** | 564 | 282 | 282 | 62 (31.8-76) | 62 (22.3-76) | 170:112<br>61% males | 168:114<br>59.5% males | 0: 139 (49%)<br>1:116 (41%)<br>2: 27 (10%) | 0:134 (47%)<br>1: 115 (41%)<br>2: 33 (12%) |
| **Garufi et al 2006** | 68 | 33 | 35 | 62 (35-77) | 61 (28-77) | 19:14<br>57.5% males | 22:13<br>62.8% males | 0: 22 (67%)<br>1: 9 (27%)<br>2: 2 (6%) | 0: 22 (63%)<br>1: 12 (34%)<br>2: 1 (3%) |
| **Focan et al 1997** | 56 | 27 | 29 | 58 (34-75) | 64 (44-75) | 17:10<br>63% males | 18:11<br>62% males | 0: 11(41%)<br>1: 14 (52%)<br>2: 2 (7%) | 0: 12 (41%)<br>1: 16 (56%)<br>2: 1 (3%) |
| **Lévi et al 1997** | 186 | 93 | 93 | 61 (29-75) | 61 (22-75) | 60:33<br>64.5% males | 52:41<br>56% males | 0: 50 (54%)<br>1: 34 (37%)<br>2-3: 9 (10%) | 0: 49 (53%)<br>1: 30 (32%)<br>2-3: 14 (15%) |
| **Lévi et al 1994** | 92 | 47 | 45 | 60 (34-73) | 60 (31-73) | 27:20<br>57.4% males | 20:25<br>44.4% males | 0: 14 (30%)<br>1:27 (57%)<br>2: 6 (13%) | 0: 19 (42%)<br>1: 23 (51%)<br>2: 3 (7%) |

Group A: Conventional (non-chronomodulated) chemotherapy – Group B: chronomodulated chemotherapy

**Supplementary table 6:** Disease Characteristics

| Study | Primary site<br>Colon: Rectum (ratio) |  | Metastasis |  | Previous chemotherapy |  | Previous radiotherapy |  | Previous Surgery |  |
| --- | --- | --- | --- | --- | --- | --- | --- | --- | --- | --- |
|  | Group A | Group B | Group A | Group B | Group A | Group B | Group A | Group B | Group A | Group B |
| **Ramanathan et al** | 76:28 (2.7) | 16:9 (1.7) | Not specified |  | Not specified |  | Not specified |  | Not specified |  |
| **Huang et al** | 14:6 (2.3) | 16:6 (2.6) | Liver metastasis > 2 cm: 17(85%). both lobes: 12 | Liver metastasis > 2 cm: 18 (81.8%)<br>Both lobes :16 | Not specified |  | Not specified |  | Not Specified |  |
| **Giacchetti et al** | 213:69 (3) | 217:65 (3.3) | 3 or more metastasis: 48 (17%) | 3 or more metastasis: 38 (13%) | 48 (17%) | 54 (19%) | 18 (6%) | 26 (9%) | Surgery for metastasis: 14 (5%) | Surgery for metastasis 14 (5%) |
| **Garufi et al** | 28:5 (5.6) | 28:7 (4) | 2 or more: 10 (30%) | 2 or more: 13 (37%) | 9 (27%) | 7 (20%) | 2 (6%) | 3 (9%) | Not specified |  |
| **Focan et al** | 18:9 (2) | 23:6 (3.8) | Isolated liver metastasis: 22 (81.4%) | Isolated liver metastasis: 23 (79.3%) | 6 (22%) | 10 (34%) | 0 (0%) | 3 (10.3%) | 27 (100%) | 29 (100%) |
| **Lévi et al 1997** | 66:27 (2.4) | 63:30 (2.1) | Less than 3: 85 (91%) | Less than 3: 85 (91%) | 11 (12%) | 10 (11%) | 7 (8%) | 6 (6%) | Surgery for metastasis: 7 (8%) | 22 (24%) |
| **Lévi et al 1994** | 30:17 (1.76) | 34:11(3.1) | 2 or more: 20 (42.5%) | 2 or more: 23 (51.1%) | 2 (4.2%) | 5 (11.1%) | 4 (8.5%) | 5 (11.1%) | Not specified |  |

Group A: Conventional (non-chronomodulated) chemotherapy – Group B: chronomodulated chemotherapy

**Supplementary table 7:** Chemotherapy Regimens

| Study | Regimen | Duration | Method of delivery | Chronomodulation strategy |
| --- | --- | --- | --- | --- |
| **Ramanathan et al** | Regimen 1: 5-FU, LV, Oxaliplatin | up to 24weeks or disease progression | IV | Only 5 FU was chronomodulated with 5 hours infusion with peak at 04:00hrs. Oxaliplatin was administered immediately before 5 FU over 6 hrs in day 1 every 3 weeks |
| **Huang et al** | Regimen 2: Arterial 5-FU and Oxaliplatin | each patient arranged to receive 2 courses then further chemotherapy or radio-ablation depending on response. | Seldinger technique to cannulate the hepatic artery proper if metastasis on both lobes. Right or left hepatic artery cannulation if confined to one lobe. Gastroduodenal artery embolisation if necessary. Porth Cath System (PCS) subcutaneous over the inguinal region. | 5 FU infused from 22:00h to 10:00h with peak flow at 4:00h. Oxaliplatin infused between 10:00h to 22:00 with peak flow at 16:00h |
| **Giacchetti et al** | Regimen 1: 5-FU, LV, Oxaliplatin | 4 days for chronomodulated and 2 days for non-chronomodulated course. Courses were repeated every 14 days | IV | 5 FU and LV from 2215 to 0945 with peak at 0400, oxaliplatin from 1015 to 2145 with peak at 1600hrs |
| **Garufi et al** | Regimen 3: Irinotecan (CPT-11) ,5-FU, LV (Both groups received chronomodulated 5-FU and LV) | 5 days course every 2 weeks, continued until progression, unacceptable toxicity, or patient refusal | IV | CPT-11 given as a 6-h sinusoidal infusion from 2:00 to 8:00 a.m. with peak flow at 5:00 a.m. in 250 ml of 5% dextrose |
| **Focan et al** | Regimen 4: Venous 5-FU, arterial FUDR | 5 days course followed by 16 days treatment free interval for at least 6 courses | All patients had staging laparotomy, cholecystectomy, and gastroduodenal artery ligation at the time of surgical placement of the catheter into the hepatic artery | peak at 04:00 for 5FU and 16:00 for FUDR |
| **Lévi et al 1997** | Regimen 1: 5-FU, LV, Oxaliplatin | Each 5-day course was repeated after a 16-day interval. | IV | Peak flow at 04:00 for 5 FU and 16:00 for Oxaliplatin |
| **Lévi et al 1994** | Regimen 5: 5-FU, I-OHP, LV | Each 5-day course was repeated after a 16-day interval for at least 6 cycles then offered surgery if response | IV | Peak at 04:00 for 5 FU and LV. Peak at 16:00 for I-OHP |

**Supplementary table 8:** Meta-regression of chemotherapy regimens

| Covariate reference | Coefficient | Standard error | 95% Lower | 95% Upper | Z-Value | 2-sided P-value | Set |
| --- | --- | --- | --- | --- | --- | --- | --- |
| **Intercept** | 0.2952 | 0.2579 | -0.2102 | 0.8007 | 1.14 | 0.2522 |  |
| **chemotherapy regimen: 2** | -2.5785 | 0.9144 | -4.3706 | -0.7864 | -2.82 | 0.0048 | Q=10.45, df=5, p=0.0635 |
| **chemotherapy regimen: 3** | -0.3154 | 0.4858 | -1.2675 | 0.6367 | -0.65 | 0.5162 | Q=10.45, df=5, p=0.0635 |
| **chemotherapy regimen: 4** | -0.1188 | 0.7673 | -1.6227 | 1.3851 | -0.15 | 0.877 | Q=10.45, df=5, p=0.0635 |
| **chemotherapy regimen: 5** | -0.5965 | 0.547 | -1.6686 | 0.4757 | -1.09 | 0.2755 | Q=10.45, df=5, p=0.0635 |
| **Statistics for Model 1** |  |  |  |  |  |  |  |
| **Test of the model: Simultaneous test that all coefficients (excluding intercept) are zero** |  |  |  |  |  |  |  |
| Q = 10.45, df = 5, p = 0.0635 |  |  |  |  |  |  |  |
| **Goodness of fit: Test that unexplained variance is zero** |  |  |  |  |  |  |  |
| Tau<sup>2</sup> = 0.6669, Tau = 0.8167, I<sup>2</sup> = 83.90%, Q = 180.12, df = 29, p = 0.0000 |  |  |  |  |  |  |  |
| **Comparison of Model 1 with the null model** |  |  |  |  |  |  |  |
| **Total between-study variance (intercept only)** |  |  |  |  |  |  |  |
| Tau<sup>2</sup> = 0.6018, Tau = 0.7758, I<sup>2</sup> = 83.97%, Q = 212.08, df = 34, p = 0.0000 |  |  |  |  |  |  |  |
| **Proportion of total between-study variance explained by Model 1** |  |  |  |  |  |  |  |
| R<sup>2</sup> analog = 0.00 (computed value is -0.11) |  |  |  |  |  |  |  |

**Supplementary figure 1:**
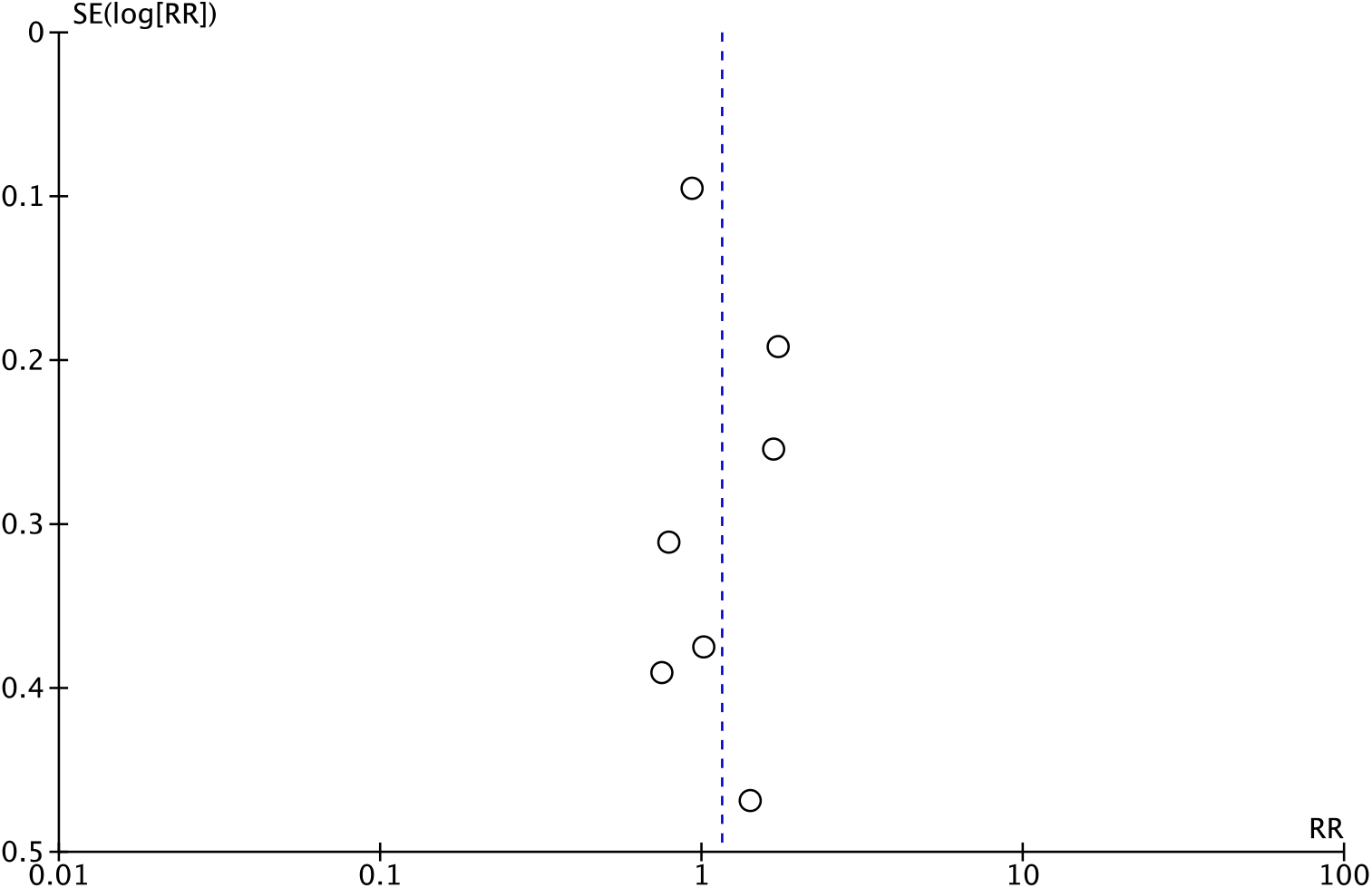
Funnel plot (Objective response rate)

**Supplementary figure 2:**
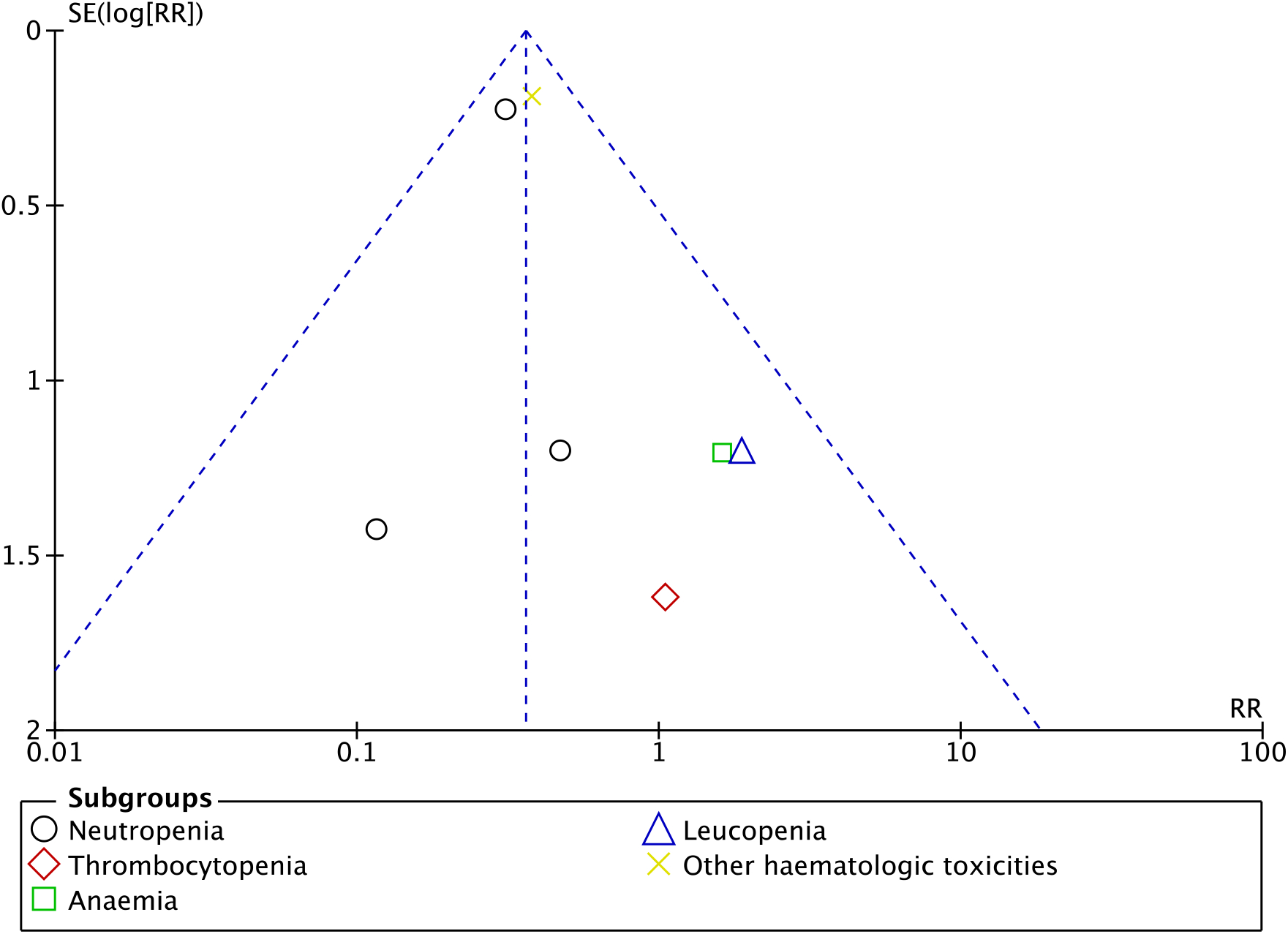
Funnel Plot (Haematological toxicity)

**Supplementary figure 3:**
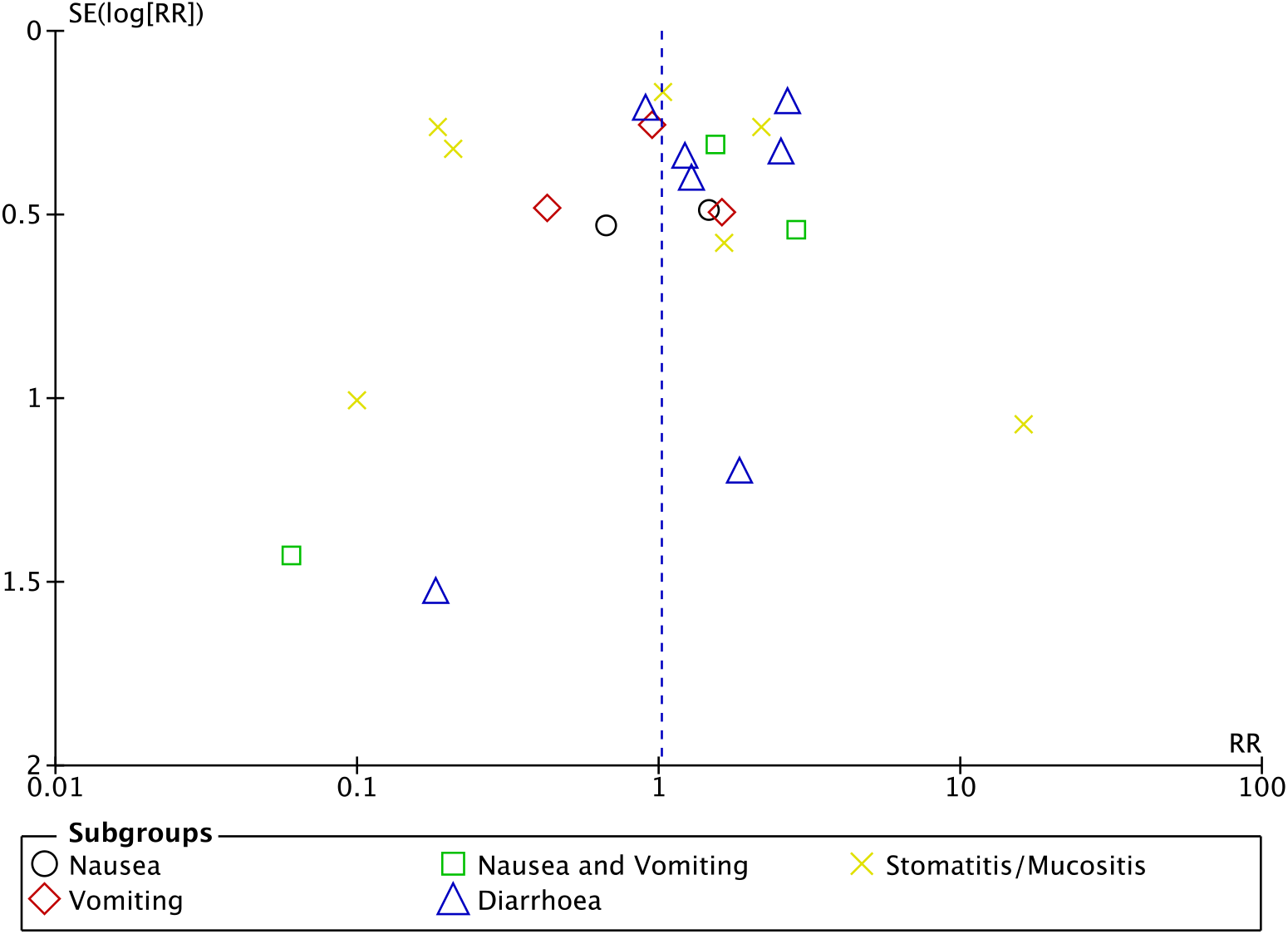
Funnel Plot (Gastrointestinal toxicity)

**Supplementary figure 4:**
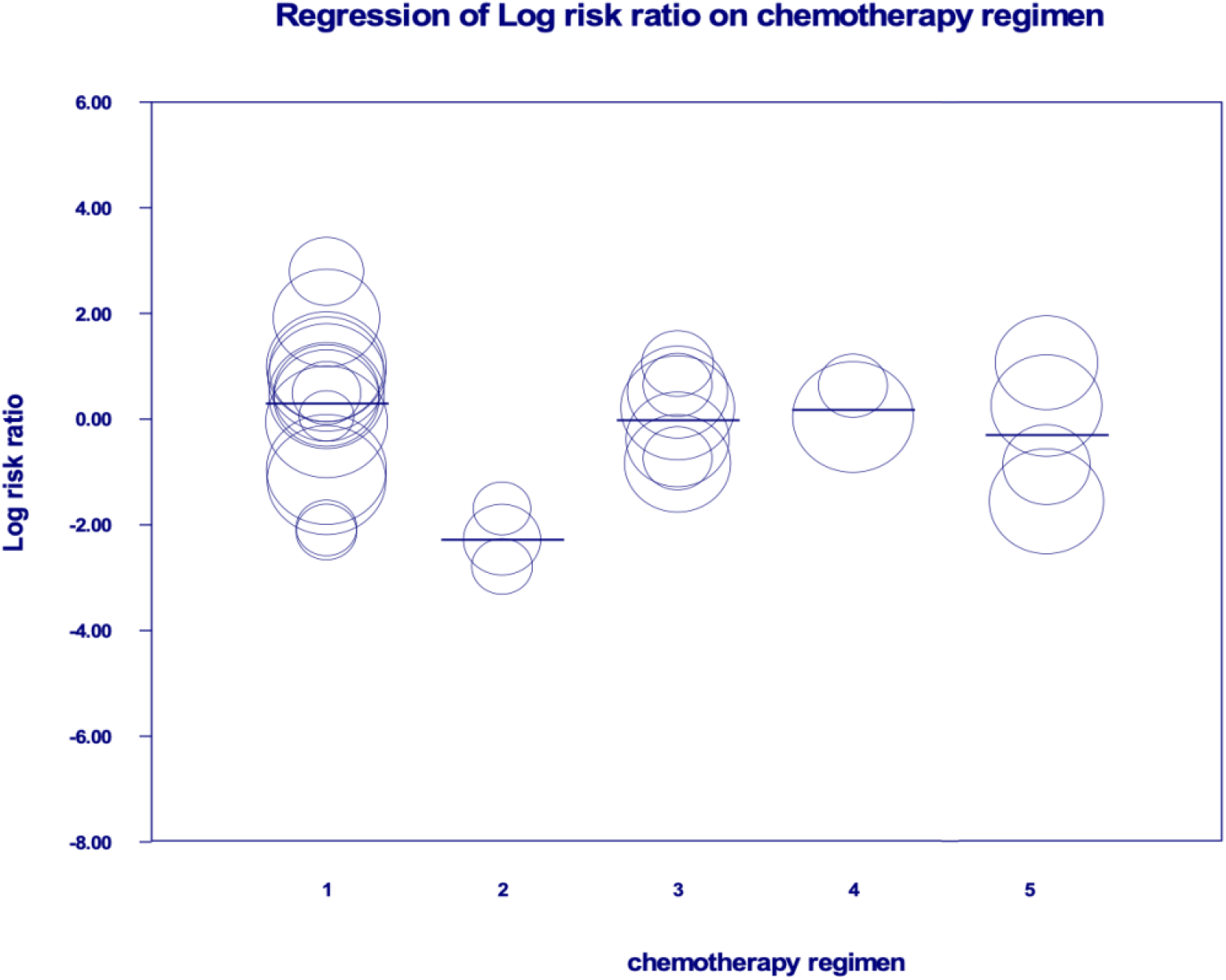
Meta-regression: chemotherapy regimen 1: 5FU, LV plus oxaliplatin; chemotherapy regimen 2: intra-arterial 5 FU and oxaliplatin; chemotherapy regimen 3: CPT-11, 5FU and LV; chemotherapy regimen 4: venous 5-FU and arterial FUDR; chemotherapy regimen 5: 5-FU, I-OHP and LV

## Notes

### Competing Interest Statement

The authors have declared no competing interest.

### Funding Statement

This study did not receive any funding

